# Impact of Intravenous Iron Therapy on Hospitalizations and Mortality in Patients with Heart Failure and Iron Deficiency: A Systematic Review and Meta-Analysis

**DOI:** 10.1101/2024.02.02.24302246

**Authors:** Haiming Wang, Yanhua Li, Jingjing Zhou, Jing Wang, Junjie Shao, Shuai Yue, Jiayue Li, Xinhong Guo, Ran Zhang

## Abstract

**BACKGROUND:** Intravenous iron therapy represents a promising potential treatment option for patients with heart failure (HF) and iron deficiency (ID), as it has been shown to improve clinical symptoms and enhance quality of life. To investigate the benefits of intravenous iron therapy on hard cardiovascular endpoints in HF and ID patients, we conducted a systematic review and meta-analysis of randomized controlled trials (RCTs).

**METHODS:** We implemented a systematical search of the PubMed, Embase and Cochrane Library databases for relevant RCTs of intravenous iron therapy in patients with HF and ID published from inception through January 20, 2024. Our primary endpoints of interest were HF hospitalizations, all-cause mortality, HF hospitalizations and cardiovascular death, cardiovascular hospitalizations and cardiovascular death. Sensitivity analyses and subgroup analyses were further performed to investigate additional clinical benefits in specific populations.

**RESULTS:** Eleven trials encompassing a collective cohort of 6511 participants met our predefined eligibility criteria and were included in our meta-analysis. The predominant form of intravenous iron utilized in the trials included in our analysis was ferric carboxymaltose. Intravenous iron therapy yielded a 40% relative reduction in HF hospitalization (OR 0.60, 95% CI 0.51-0.70; *P*= 0.00001), a 46% relative reduction in HF hospitalizations and cardiovascular death (OR 0.54, 95% CI 0.46-0.63; *P*<0.00001) and a 53% relative reduction in cardiovascular hospitalizations and cardiovascular death (OR 0.47, 95% CI 0.37-0.59; *P*<0.00001). Our analysis revealed no statistically significant differences in terms of all-cause mortality (OR 0.85, 95% CI 0.72-1.01; *P*=0.06) while this result was fragile (reverse fragility index of 2 and reverse fragility quotient of 0.0004). Subgroup analyses revealed more favorable effects of intravenous iron therapy in trials that had a follow-up duration of ≥ 24 weeks and a sample size of over 200 cases. Intravenous iron therapy had negligible effects on infection (OR 0.86, 95% CI 0.66-1.11; *P*=0.25), general disorders and administration site conditions (OR 1.35, 95% CI 0.93-1.94; *P*=0.11), injury, poisoning and procedural complications (OR 0.96, 95% CI 0.66-1.40; *P*=0.85).

**CONCLUSION:** Intravenous iron therapy in patients with HF and ID shows a significant reduction of rehospitalization for HF and cardiovascular death. The ferric carboxymaltose holds significant promise as a potential therapeutic agent for HF patients with ID.

## INTRODUCTION

Heart failure (HF) is a chronic and progressive syndrome that severely compromises the quality of life and significantly reduces the life expectancy ^1^. Despite great advances in drug and device therapy for HF, the mortality and rehospitalization rates remain alarmingly high ^2–4^. Iron deficiency (ID) is a highly prevalent comorbidity in patients with HF, causing the aggravation of debilitating symptoms, decreased exercise capacity, elevated rehospitalization rates, and heightened mortality risk ^5–9^. However, the routine screening and standard therapy for ID have not been widely integrated into the current management of patients with HF ^2, 5^. Intravenous iron repletion has been shown to improve clinical symptoms, enhance the 6-minute walk distance, and potentially reduce the risk of rehospitalization in patients with HF and ID ^10–13^. Therefore, the administration of intravenous iron has been recommended for patients with HF who have ID and a left ventricular ejection fraction (LVEF) below 50% in order to improve symptoms, enhance their quality of life and reduce hospitalizations related to HF ^5, 14^. However, the recent HEART-FID trial (Ferric Carboxymaltose in Heart Failure with Iron Deficiency) has revealed minimal impact of intravenous iron on significant cardiovascular outcomes, such as mortality and HF-related hospitalizations, among ambulatory patients with both HF and ID ^15^. To assess the efficacy of intravenous iron therapy on cardiovascular outcomes in patients with HF and ID, we conducted a systematic review and meta-analysis that incorporated existing clinical evidence from published RCTs.

## MATERIALS AND METHODS

### Search Strategy and Selection Criteria

The present systematic review and meta-analysis adhered to the guidelines set forth by the Preferred Reporting Items for Systematic Reviews and Meta-Analyses (PRISMA) statement^16^. We systematically searched the PubMed, Embase, and Cochrane Library databases for relevant RCTs investigating the efficacy of intravenous iron therapy in patients with HF and ID, covering the period from inception through January 20, 2024, without imposing any language restrictions. We used the following MeSH terms and combined text to guide our search: "heart failure" and "iron compounds" or "ferric carboxymaltose" or "iron deficiency" or "iron therapy" or "iron derivative" or "iron saccharate". In addition, we manually searched the references of relevant systematic reviews and meta-analyses to ensure inclusion of all eligible studies. We did not impose any restrictions on the number of patients, type of intravenous iron, or length of follow-up in potentially eligible studies.

### Study Selection and Data Extraction

We included RCTs that compared intravenous iron therapy to placebo in HF patients with ID. All eligible studies had to report at least one of the predefined clinical primary endpoints, including HF hospitalizations, all-cause mortality, HF hospitalizations and cardiovascular death, or cardiovascular hospitalizations and cardiovascular death. The secondary clinical endpoints encompassed time to first hospitalization, time to first cardiovascular hospitalization, time to first cardiovascular death, time to first HF hospitalization or cardiovascular death, time to first hospitalization or all-cause mortality, and time to first cardiovascular hospitalization or cardiovascular death. We also assessed the safety of intravenous iron by collecting data on adverse effects. Exclusion criteria comprised retrospective or observational trials, RCTs that utilized oral iron therapy, and RCTs that did not report our predetermined outcomes of interest.

Following the database searches, two independent investigators (HW and YHL) screened the retrieved studies based on their titles and abstracts. The studies that met the inclusion criteria were then evaluated in full-text form. Two investigators (HW and YHL) extracted data and conducted comprehensive analysis, while a third investigator (RZ) acted as an arbitrator in cases of disagreement. The quality of the studies was assessed according to PRISMA recommendations, and the risk of bias was evaluated using the Cochrane Risk of Bias Tool.

### Statistical Analysis

We evaluated ten categorical variable outcomes to assess the clinical benefits of intravenous iron therapy. Given the heterogeneity of all eligible trials and its potential influences on beneficial effects, we used the Cochran *Q* test and *I²* test to evaluate the magnitude of the heterogeneity between trials for each outcome. When *P* value was less than 0.1 or the value of the *I²* test was greater than 50%, the evaluation of outcome exhibited significant heterogeneity. In cases where high heterogeneity was observed, the clinical effects were evaluated using a random-effects model based on the DerSimonian–Laird method. Alternatively, if heterogeneity was not significant, a fixed-effects model based on the Mantel–Haenszel method was utilized. Pooled estimates were presented as odds ratios (ORs) with corresponding 95 percent confidence intervals (CIs). Sensitivity analyses using the leave-one-out method were conducted to identify any specific trial that contributed to greater heterogeneity. The effect sizes of the remaining trials were recalculated to assess the robustness and reliability of the findings.

To further assess the robustness of the meta-analysis results, we employed the Fragility Index (FI) for statistically significant findings and the Reverse Fragility Index (RFI) for non-significant findings. These indices helped determine the vulnerability or stability of the results and provided additional insights into the reliability of the findings^17, 18^. The FI or RFI was calculated as the minimum number of patients in one or more trials included in the meta-analysis, where a modification to the event status (i.e., changing an event to a non-event or vice versa) would alter the statistical significance of the pooled estimates. By identifying the minimum number of patients required to influence the statistical significance, these indices provided valuable information on the fragility of the results and their susceptibility to change^18^. FI or RFI <20 was considered as a fragile outcome, and FI or RFI ≥40 as a robust outcome^17, 18^. We combine it with the sample size to calculate the fragility quotient (FQ) and reversed FQ (RFQ) that can more accurately represent the proportion of events required to alter the effect sizes^17^. For instance, Meta-analysis A has an FI of 1 and a sample size of 1,000, whereas Meta-analysis B has an FI of 1 and a sample size of 10,000. Although both analyses had the same FI, the FQ for Meta-analysis A was 0.001, indicating that it took 10 events per 10,000 patients to change the significance of the results, whereas the FQ for Meta-analysis B was 0.0001, indicating that it only took 1 event per 10,000 patients to change the significance of the results^17^. Therefore, the determination of FQ can help interpret the robustness of results.

Furthermore, a funnel plot was constructed to assess potential publication bias for each outcome, but this analysis was conducted only when the number of included studies exceeded 10. Additionally, we conducted three subgroup analyses based on the length of follow-up, number of patients, or type of intravenous iron to explore potential clinical benefits associated with these variables. To perform statistical analysis, we utilized RevMan 5.3 and Stata 16.0 software, ensuring accurate and reliable processing of the data in our meta-analysis.

## RESULTS

### Overview of the Literature Search and Study Characteristics

The initial literature search yielded a total of 469 relevant articles. After applying our predefined eligibility criteria, 11 trials comprising 6511 participants were deemed suitable for inclusion in the present meta-analysis (Figure 1). The risk of bias assessment for this meta-analysis is presented in Supplemental Figure 1. All selected studies demonstrated an overall low risk of bias, with only one study showing potential performance bias. Table 1 provides a comprehensive summary of the demographic characteristics, concomitant diseases, and medication combinations in the 11 eligible RCTs. Among all the trials that reported on intravenous iron therapy, only one trial (the PRACTICE-ASIA-HF trial) enrolled patients with acute HF, regardless of their left ventricular ejection fraction (LVEF). In contrast, the rest of the trials focused on individuals with chronic HF and a LVEF of ≤50%. ID was generally defined as a serum ferritin level <100 μg/L, regardless of transferrin saturation (TSAT), or a TSAT <20% if the ferritin level ranged from 100∼300 μg/L. Among the 11 studies included in our analysis, intravenous administration of ferric carboxymaltose was used in 8 of them. The age of the included patients ranged from 40 to 82 years old, and the duration of follow-up varied from 3 weeks to 2.7 years. Most patients had such comorbidities as coronary heart disease, diabetes, hypertension, and dyslipidemia.

**Figure 1.**
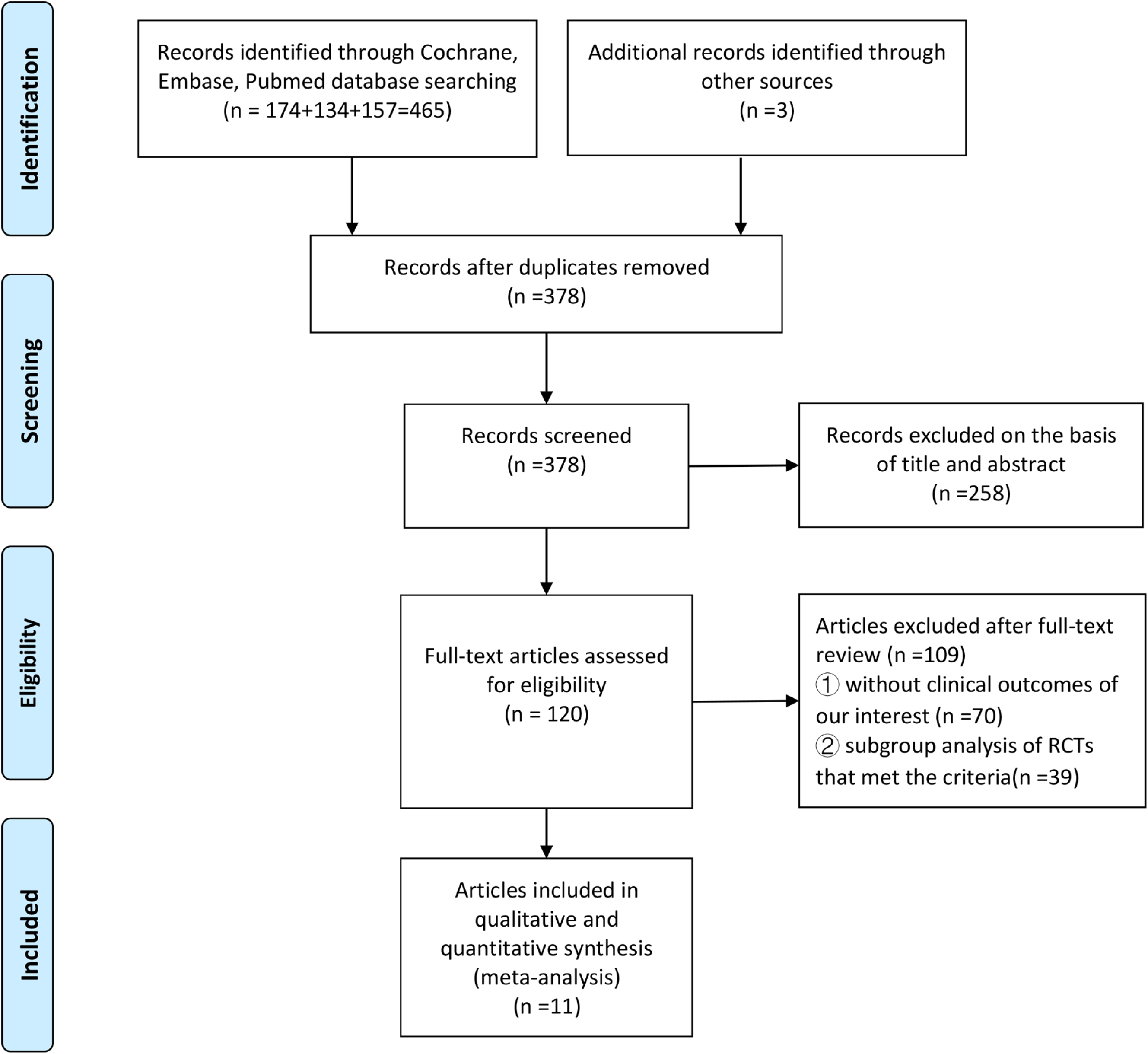
Study selection process for this systematic review

**Table 1.**
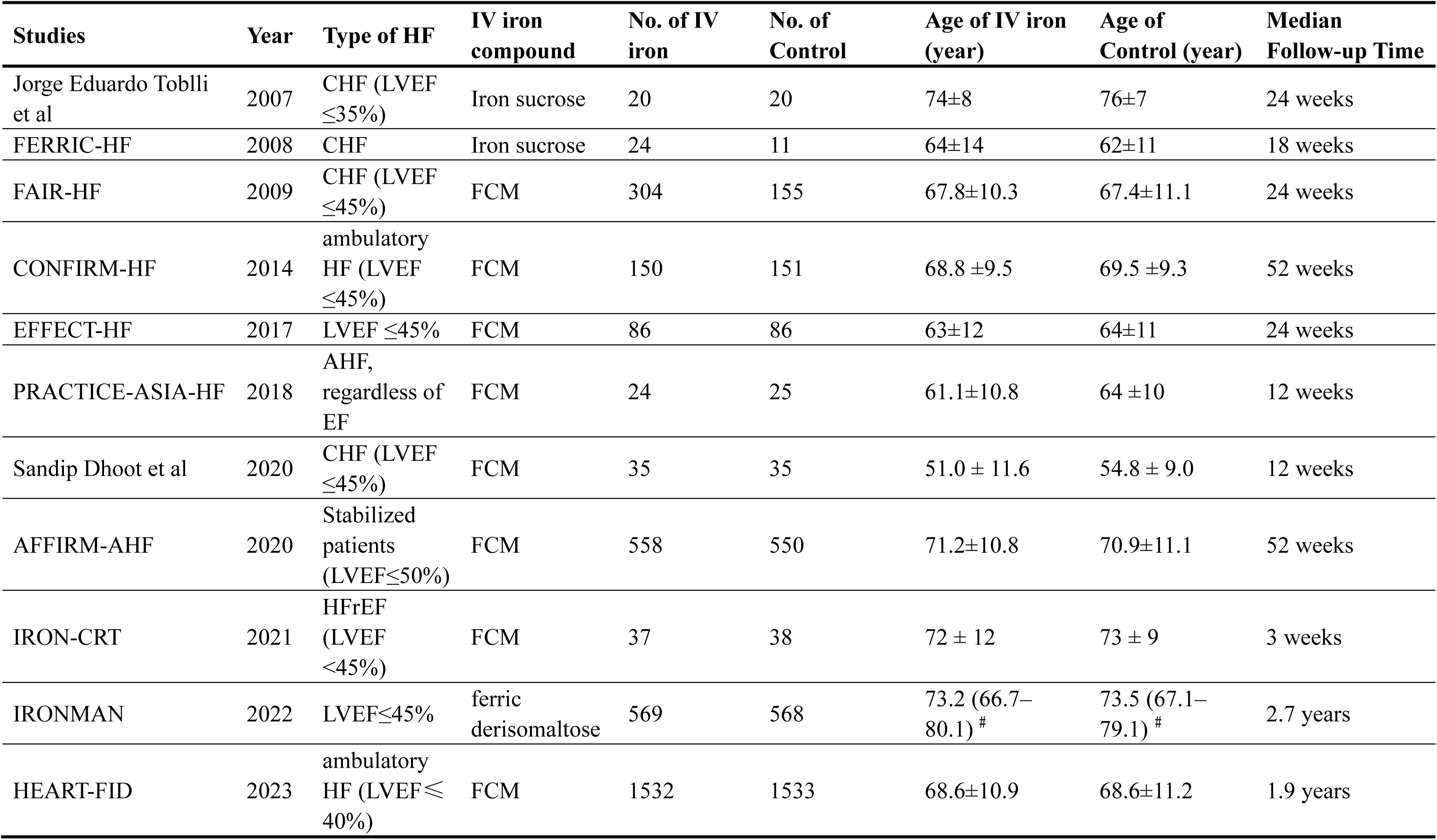
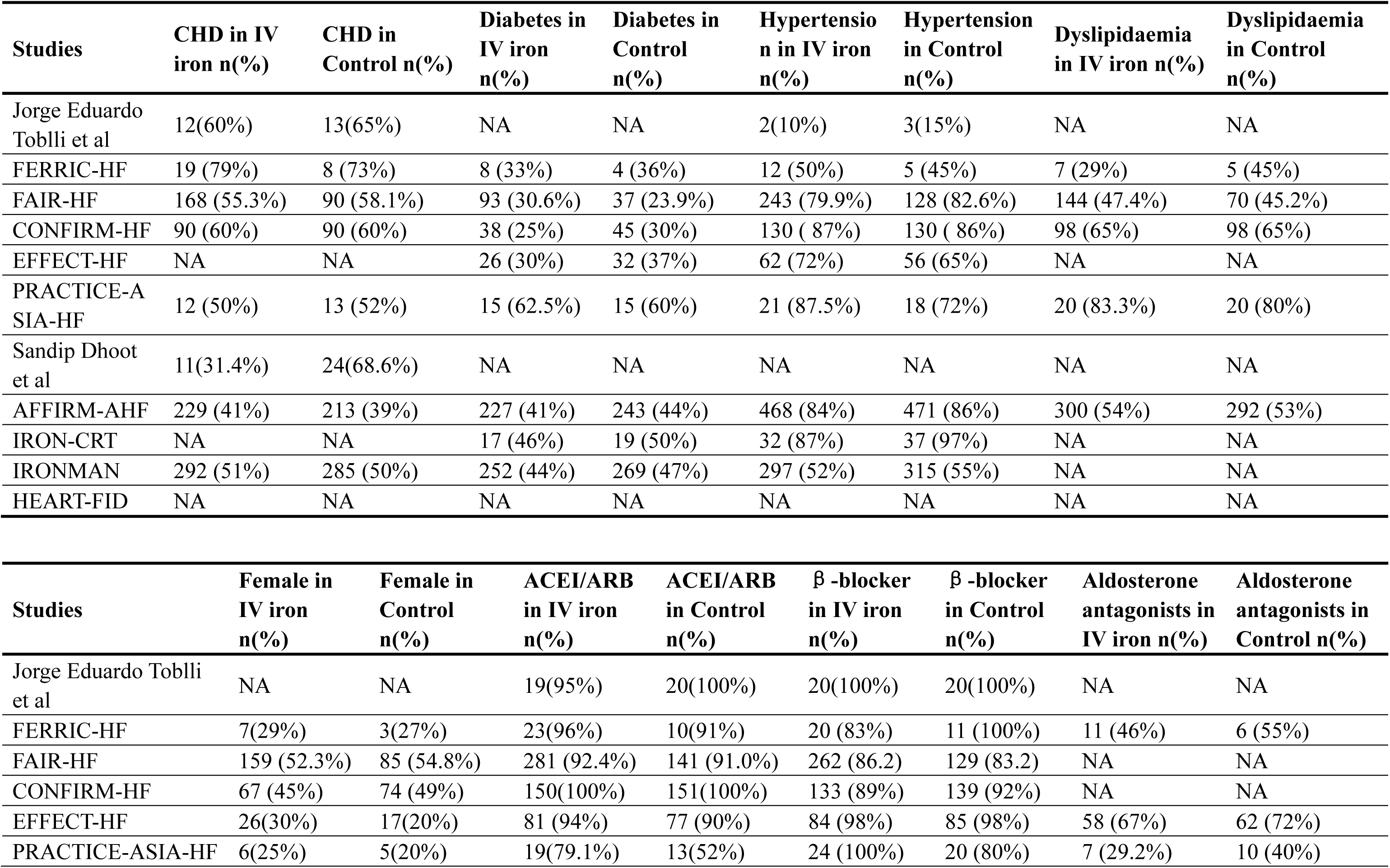
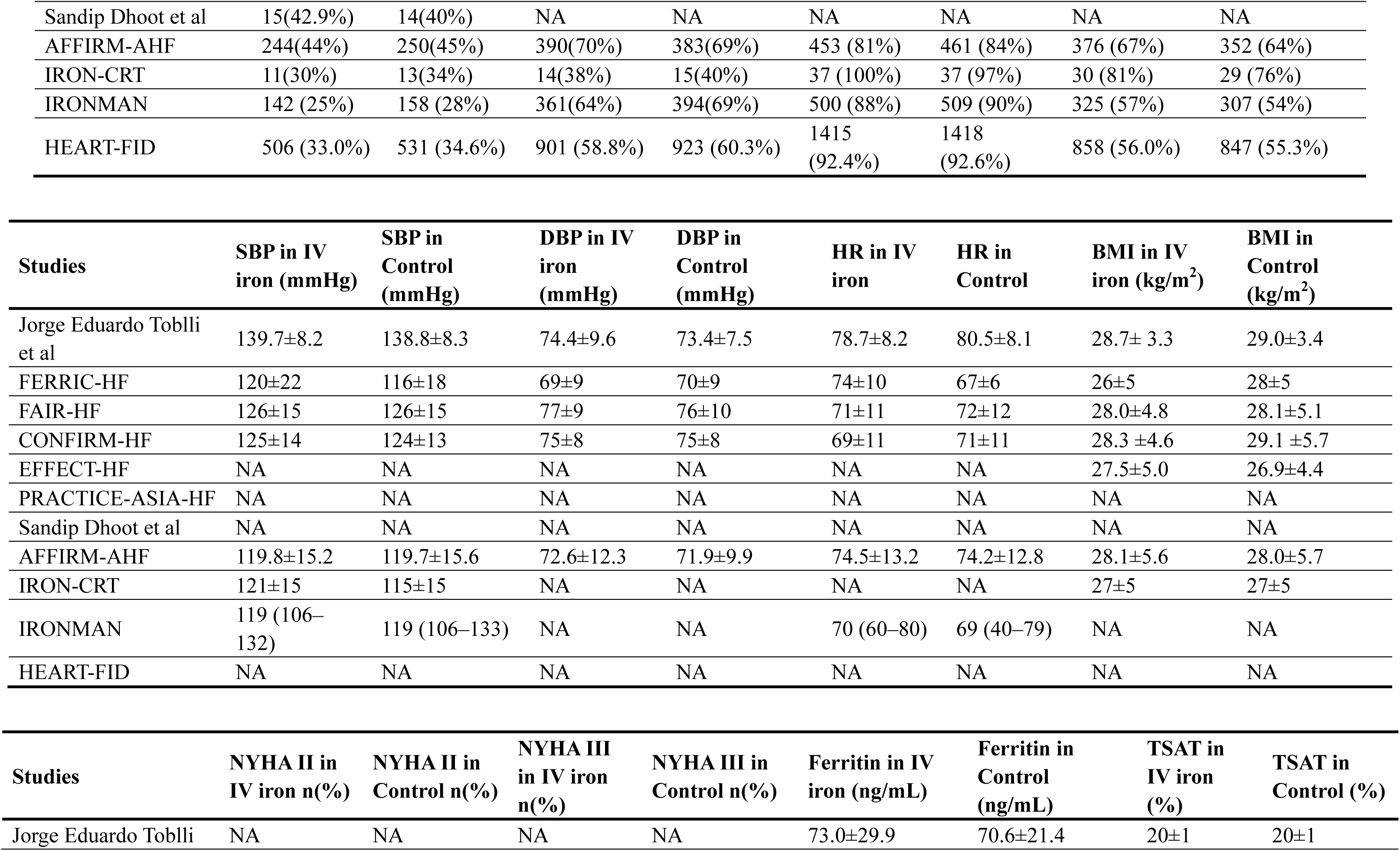
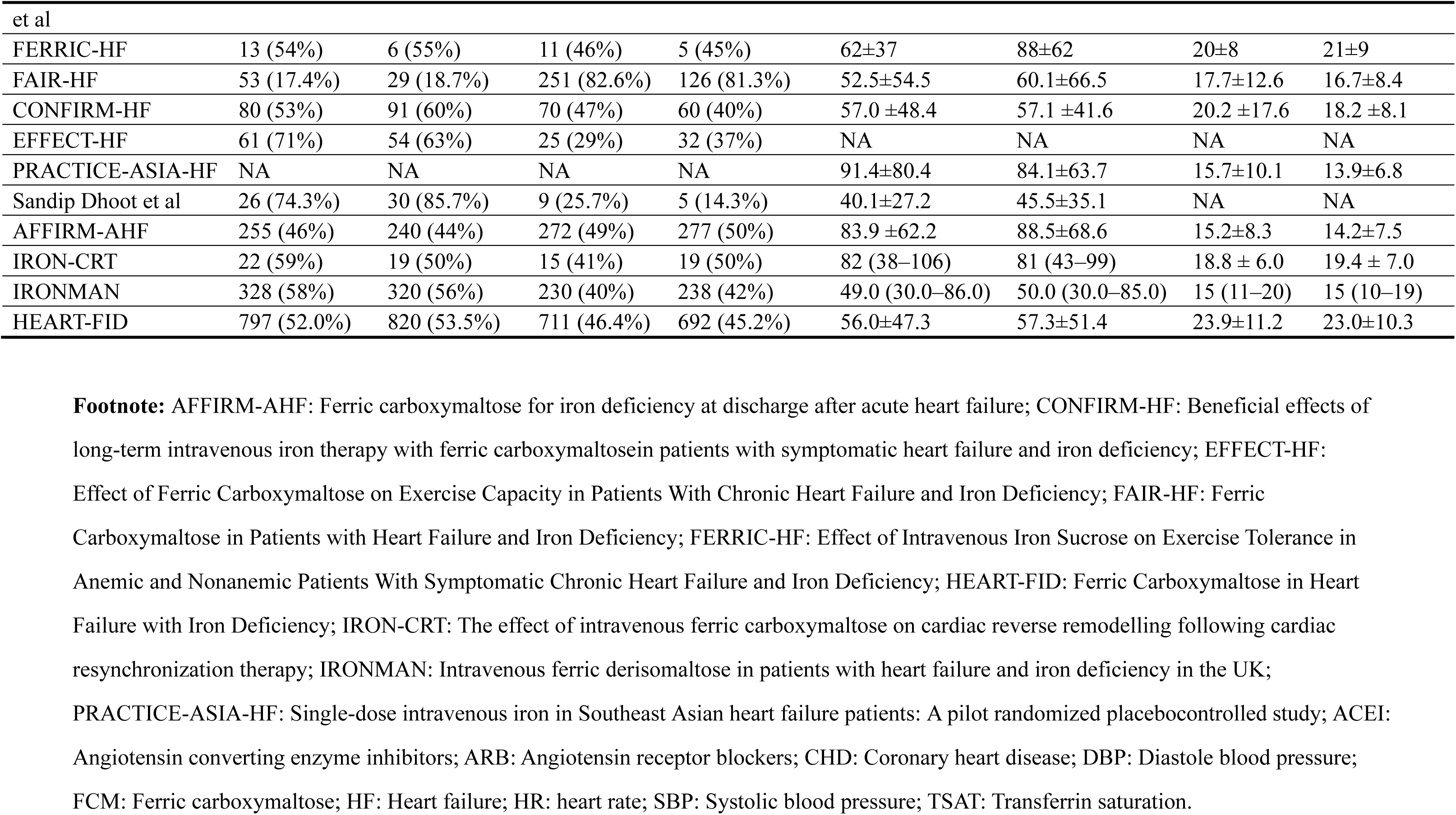
Characteristics of Included Studies.

### Primary Endpoints

Intravenous iron therapy demonstrated promising results in reducing hospitalizations related to HF (OR 0.67, 95% CI 0.51-0.88; *P*= 0.004; *I*^2^= 61%), HF hospitalizations and cardiovascular death (OR 0.62, 95% CI 0.45-0.85; *P*<0.003; *I*^2^=76%), cardiovascular hospitalizations and cardiovascular death (OR 0.58, 95% CI 0.37-0.92; *P*<0.0001; *I*^2^= 84%), with different degrees of significant between-study heterogeneities (Figure 2A, Figure 3A, Figure 4A). The calculations of FIs and FQs similarly showed these results to be fragile (Table 2 and Supplemental Figure 2-4). Through additional rigorous sensitivity analyses, heterogeneities were significantly minimized, revealing that intravenous iron treatment was associated with a substantial 40% relative reduction in HF hospitalizations (OR 0.60, 95% CI 0.51-0.70; *P* = 0.00001; *I*^2^ =36%), a 46% relative reduction in HF hospitalizations and cardiovascular death (OR 0.54, 95% CI 0.46-0.63; *P*<0.00001; *I*^2^=0%) and a 53% relative reduction in cardiovascular hospitalizations and cardiovascular death (OR 0.47, 95% CI 0.37-0.59; *P* < 0.00001; *I*^2^ = 0%), with low heterogeneities after exclusion from the HEART-FID trial (Figure 2B, Figure 3B, Figure 4B). These pooled results with higher FI and FQ were robust (Table 2 and Supplemental Figure 5-7).

**Figure 2.**
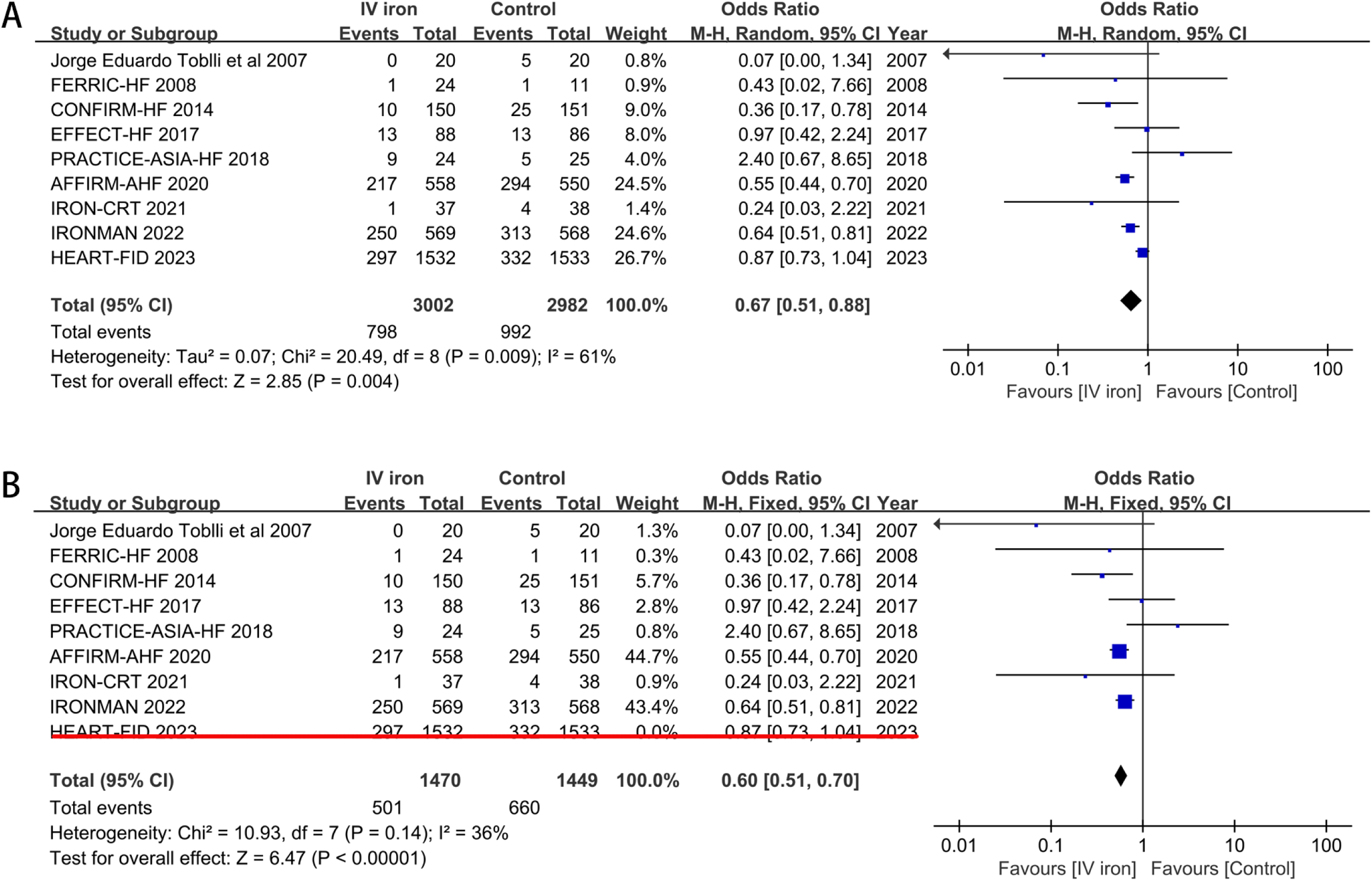
Forest plots of studies evaluating hospitalizations for heart failure in patients receiving intravenous iron therapy compared to those who received a placebo.

**Figure 3.**
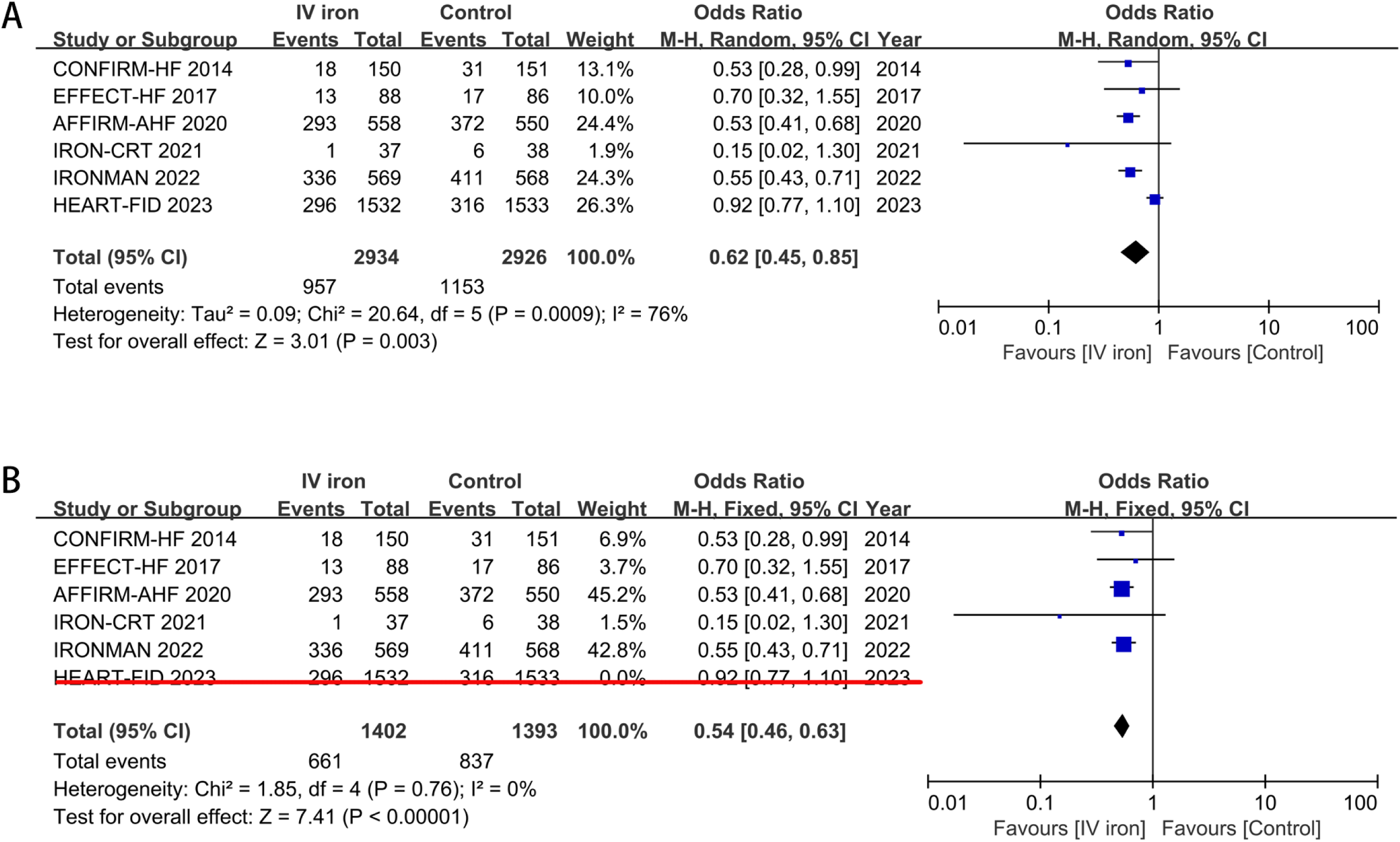
Forest plots of studies assessing HF hospitalizations and cardiovascular death in patients receiving intravenous iron therapy compared to those who received a placebo.

**Figure 4.**
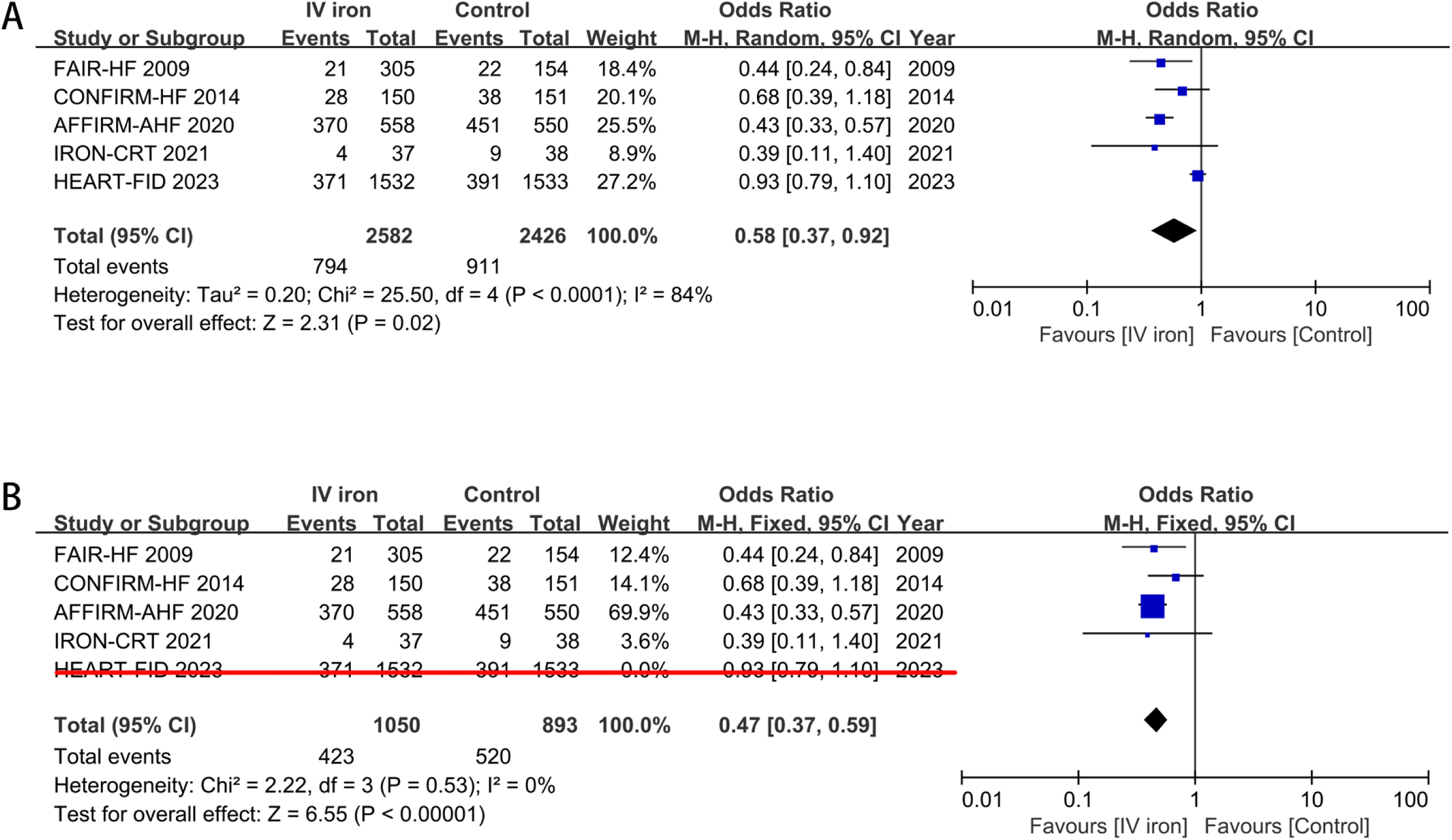
Forest plots of studies assessing cardiovascular hospitalizations and cardiovascular death in patients receiving intravenous iron therapy compared to those who received a placebo.

**Figure 5.**
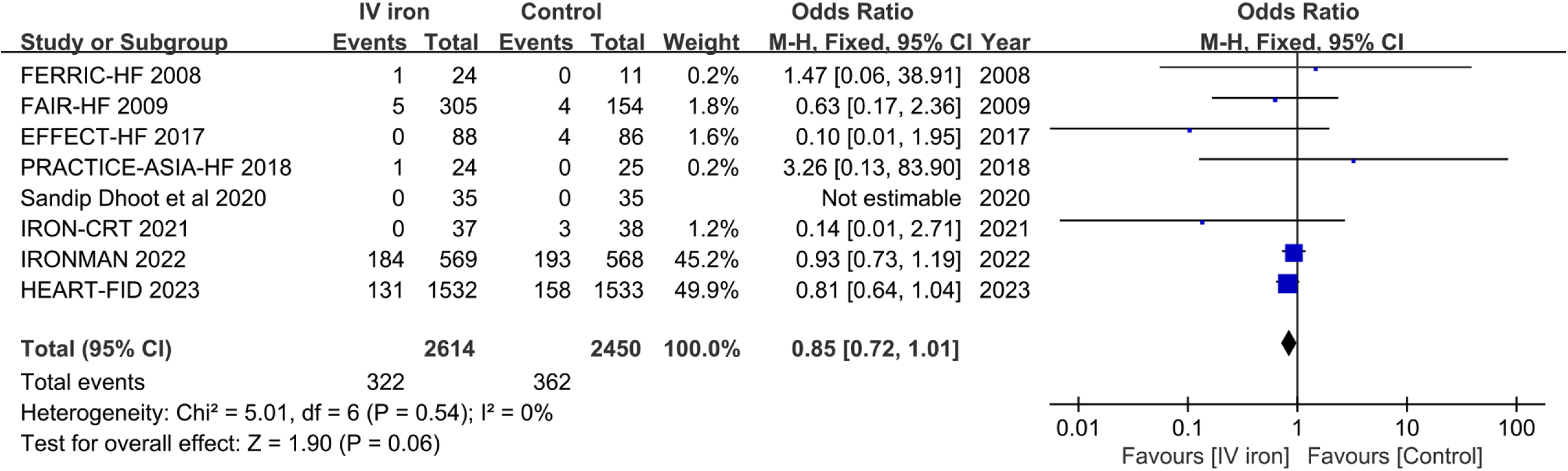
Forest plots of studies assessing all-cause mortality in patients receiving intravenous iron therapy compared to those who received a placebo.

**Table 2.** The robustness of the meta-analysis findings for clinical primary endpoints.

| Clinical primary endpoints | $I^2$ value | FI/RFI | FQ/RFQ |
| --- | --- | --- | --- |
| HF hospitalizations | 61% | 8 | 0.0013 |
| HF hospitalizations (except HEART-FID) | 36% | 111 | 0.0380 |
| HF hospitalizations and CV death | 76% | 14 | 0.0024 |
| HF hospitalizations and CV death (except HEART-FID) | 0 | 130 | 0.0465 |
| CV hospitalizations and CV death | 84% | 5 | 0.0010 |
| CV hospitalizations and CV death (except HEART-FID) | 0 | 74 | 0.0381 |
| All-cause mortality | 0 | 2 | 0.0004 |

**Footnote:** HF: Heart failure; CV: Cardiovascular; FI: Fragility index; RFI: Reversed fragility index; FQ: Fragility quotient; RFQ: Reversed FQ

In a summary analysis of 8 trials, intravenous iron therapy exhibited a notable trend towards reducing all-cause mortality, with no evidence of heterogeneity among the studies (OR 0.85, CI 0.72-1.01; *P* = 0.06; *I*^2^ = 0%; Figure 3). Despite not achieving statistical significance, this summary outcome implied fragility, as demonstrated by the RFI of 2 and the RFQ of 0.0004 (Table 2 and Supplemental Figure 8).

### Secondary Endpoints

Intravenous iron therapy demonstrated a substantial reduction in time to first hospitalization (OR 0.65, 95% CI 0.44-0.97; *P*=0.04; *I*^2^=53%), time to first HF hospitalizations or cardiovascular death (OR 0.76, 95% CI 0.61-0.95; *P*=0.01; *I*^2^=50%), time to first hospitalizations or all-cause mortality (OR 0.71, 95% CI 0.58-0.87; *P* =0.0009; *I*^2^ =37%), with different degrees of significant between-study heterogeneities. The analyses conducted did not reveal any significant differences in terms of time to first cardiovascular hospitalizations, time to first cardiovascular death, and time to first occurrence of either cardiovascular hospitalizations or cardiovascular death. Additional sensitivity analyses revealed that intravenous iron treatment enhanced the beneficial effects related to time to first hospitalization (OR 0.52, 95% CI 0.36-0.75; *P*=0.0004; *I*^2^=2%) and time to first cardiovascular hospitalizations (OR 0.41, 95% CI 0.27-0.64; *P*=0.0001; *I*^2^=0%) after exclusion from the IRONMAN trial, and time to first HF hospitalizations or cardiovascular death (OR 0.73, 95% CI 0.62-0.85; *P*=0.0001; *I*^2^=37%) after exclusion from the HEART-FID trial (Table 3 and Supplemental Figure 9-17).

**Table 3.** Time-to-first-event endpoint of the included patients with HF and ID.

| | Clinical endpoint | No. of<br>Included study | $I^2$ value | IV iron group | | Control group | | OR (95% CI) | *P* value |
| --- | --- | --- | --- | --- | --- | --- | --- | --- | --- |
|  |  |  |  | Events | Total | Events | Total |  |  |
| 1 | First hospitalization | 4 | 53% | 425 | 1044 | 466 | 893 | 0.65(0.44-0.97) | 0.04 |
|  | First hospitalization<br>(IRONMAN removed) * | 3 | 2% | 74 | 475 | 96 | 325 | 0.52(0.36-0.75) | 0.0004 |
| 2 | First CV hospitalizations | 3 | 78% | 296 | 1024 | 342 | 873 | 0.56(0.31-1.01) | 0.05 |
|  | First CV hospitalizations<br>(IRONMAN removed) * | 2 | 0 | 42 | 455 | 69 | 305 | 0.41(0.27-0.64) | 0.0001 |
| 3 | First HF hospitalization or CV death | 10 | 50% | 906 | 3305 | 1012 | 3133 | 0.76(0.61-0.95) | 0.01 |
|  | First HF hospitalization or CV death<br>(HEART-FID removed) * | 9 | 37% | 431 | 1773 | 518 | 1600 | 0.73(0.62-0.85) | 0.0001 |
| 4 | First CV death | 4 | 0 | 458 | 2809 | 503 | 2802 | 0.89(0.77-1.02) | 0.10 |
| 5 | First hospitalizations or all-cause<br>mortality | 3 | 37% | 456 | 1024 | 501 | 873 | 0.71(0.58-0.87) | 0.0009 |
| 6 | First CV hospitalization or CV death | 3 | 77% | 816 | 2251 | 901 | 2252 | 0.75(0.54-1.02) | 0.07 |

**Footnote:** HF: Heart failure; CV: Cardiovascular; ID: Iron deficiency; \* denote evaluating the overall estimate of selected endpoint for the remaining studies.

### Subgroup Analyses

There were some indications of potential differences by the length of follow-up, indicating more favorable effects of intravenous iron therapy on HF hospitalizations (OR 0.78, 95% CI 0.68-0.90; *P*=0.0006; *I*^2^=54%), HF hospitalizations and cardiovascular death (OR 0.64, 95% CI 0.47-0.87; *P*=0.004; *I*^2^=79%), cardiovascular hospitalizations and cardiovascular death (OR 0.60, 95% CI 0.37-0.98; *P*=0.04; *I*^2^=88%) in patients with a duration of follow-up ≥ 24 weeks (Figure 6).

**Figure 6.**
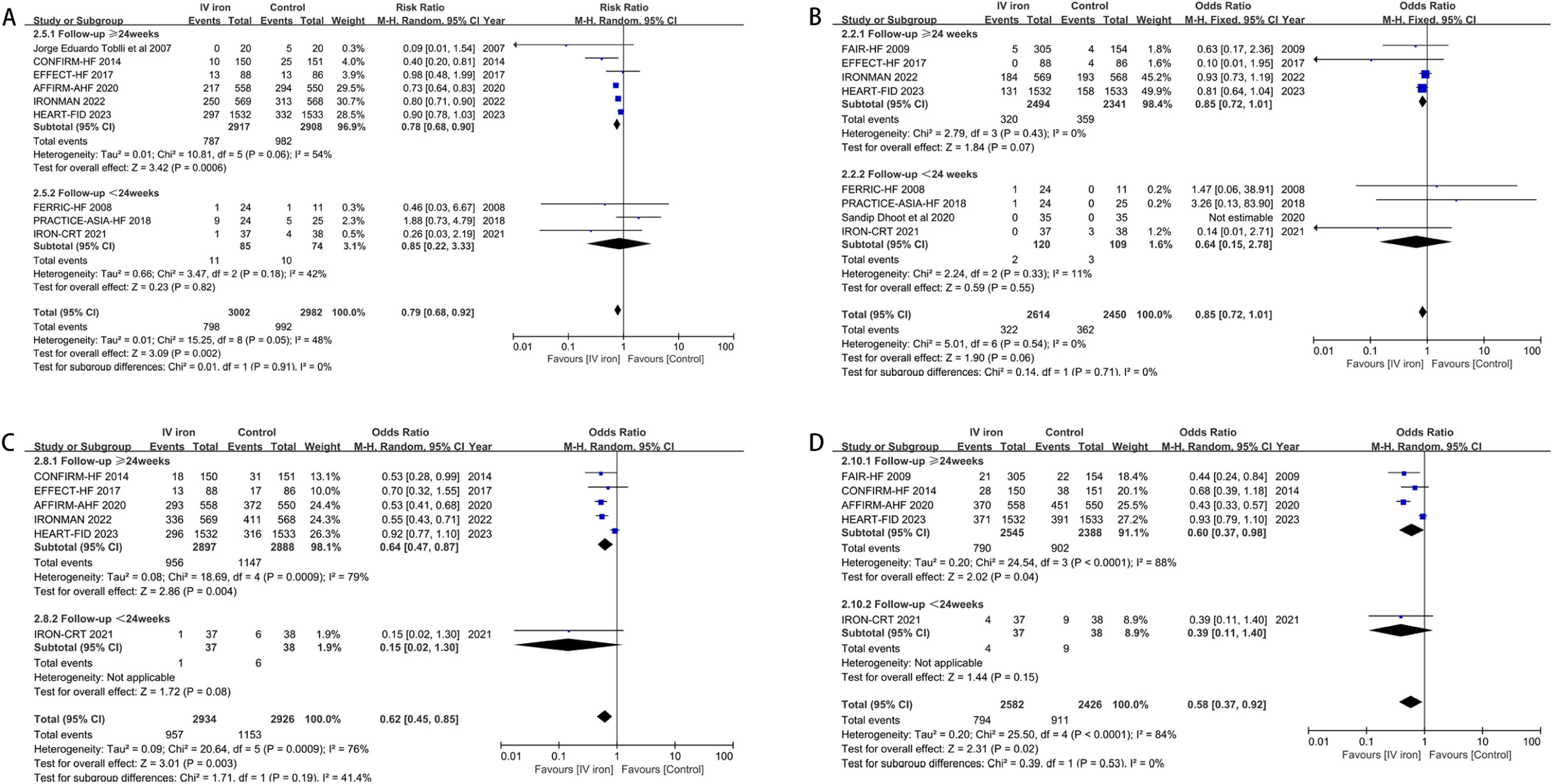
Forest plots of studies assessing primary endpoints in subgroups categorized based on varying 24-week follow-up periods.

Clinical benefits of intravenous iron therapy on HF hospitalizations (OR 0.64, 95% CI 0.48-0.85; *P*=0.002; *I*^2^=77%), HF hospitalizations and cardiovascular death (OR 0.63, 95% CI 0.45-0.88; *P*=0.008; *I*^2^=84%), cardiovascular hospitalizations and cardiovascular death (OR 0.45, 95% CI 0.38-0.53; *P*<0.00001; *I*^2^ = 0%) were equally observed when considering that the number of individuals in the study exceeded 200 cases. Likewise, the impact of treatment on all-cause mortality remained consistent regardless of the size of the sample (Figure 7).

**Figure 7.**
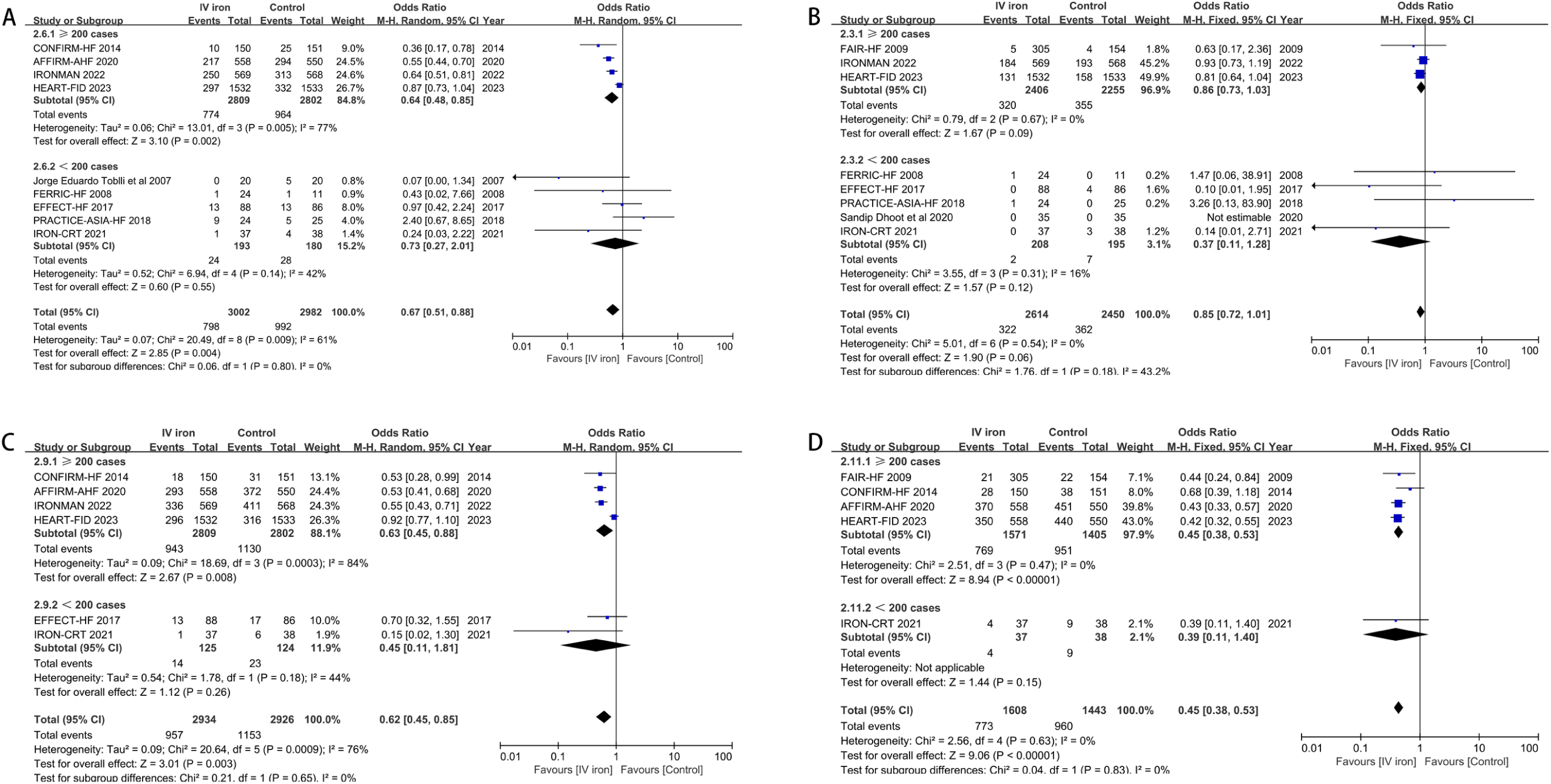
Forest plots of studies assessing primary endpoints in subgroups categorized based on a sample size of 200 patients.

The intravenous ferric carboxymaltose exerted a greater reduction in all-cause mortality (OR 0.78, 95% CI 0.62-0.99; *P*=0.04; *I*^2^=2%), HF hospitalizations and cardiovascular death (OR 0.52, 95% CI 0.44-0.61; *P*<0.00001; *I*^2^=0%), cardiovascular hospitalizations and cardiovascular death (OR 0.58, 95% CI 0.37-0.92; *P*=0.02; *I*^2^=84%) on the basis of a fixed effect model, and tended to reduce HF hospitalization rates (OR 0.70, CI 0.48-1.03; *P*=0.07; *I*^2^ = 70%) (Figure 8).

**Figure 8.**
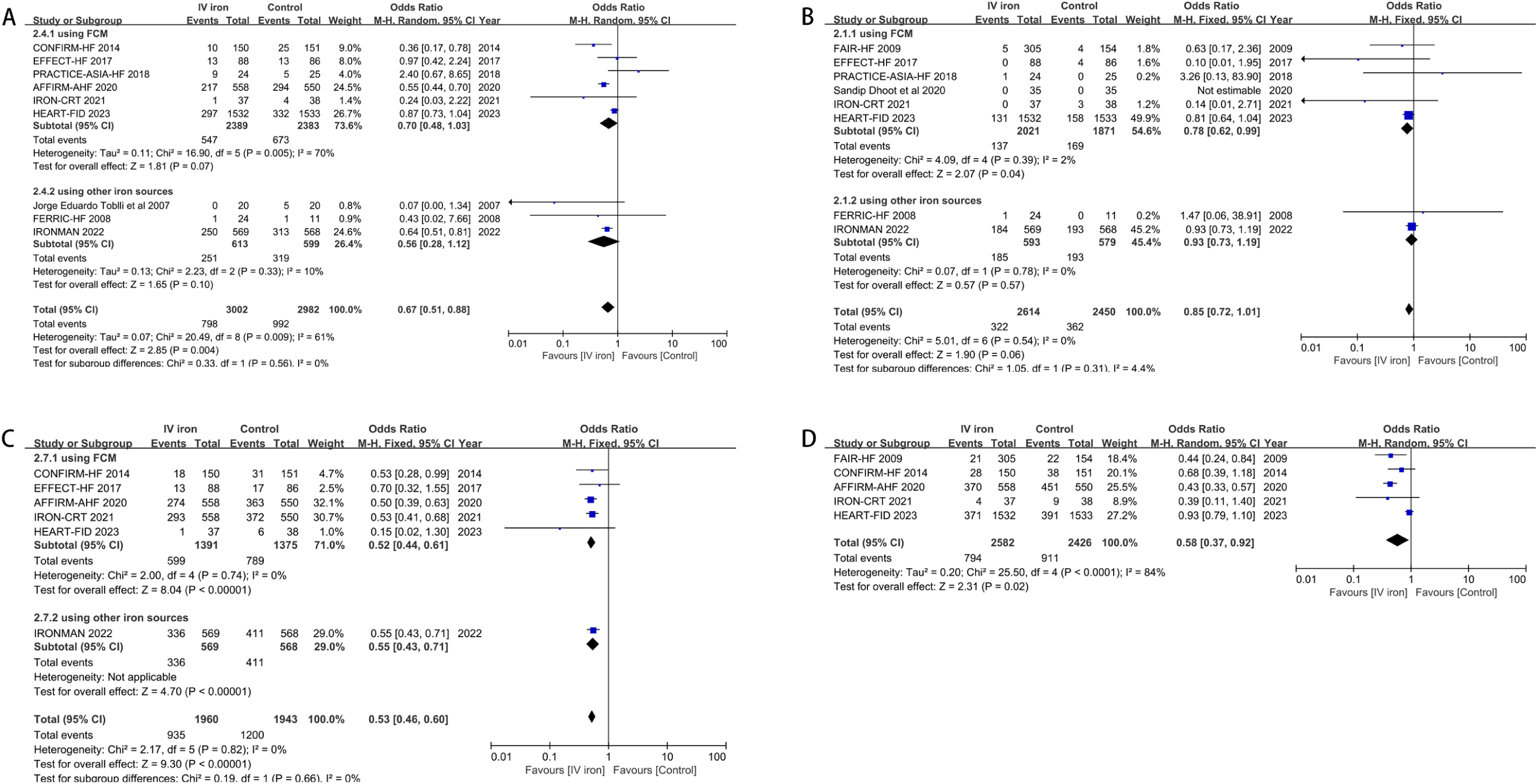
Forest plots of studies assessing primary endpoints in subgroups categorized based on the type of intravenous iron agent administered.

### Safety Endpoints

The administration of intravenous iron had minimal impacts on infection (OR 0.86, 95% CI 0.66-1.11; *P*=0.25; *I*^2^=0%), general disorders and administration site conditions (OR 1.35, 95% CI 0.93-1.94; *P*=0.11; *I*^2^=38%), injury, poisoning and procedural complications (OR 0.96, 95% CI 0.66-1.40; *P*=0.85; *I*^2^=0%), with no significant between-study heterogeneity (Figure 9).

**Figure 9.**
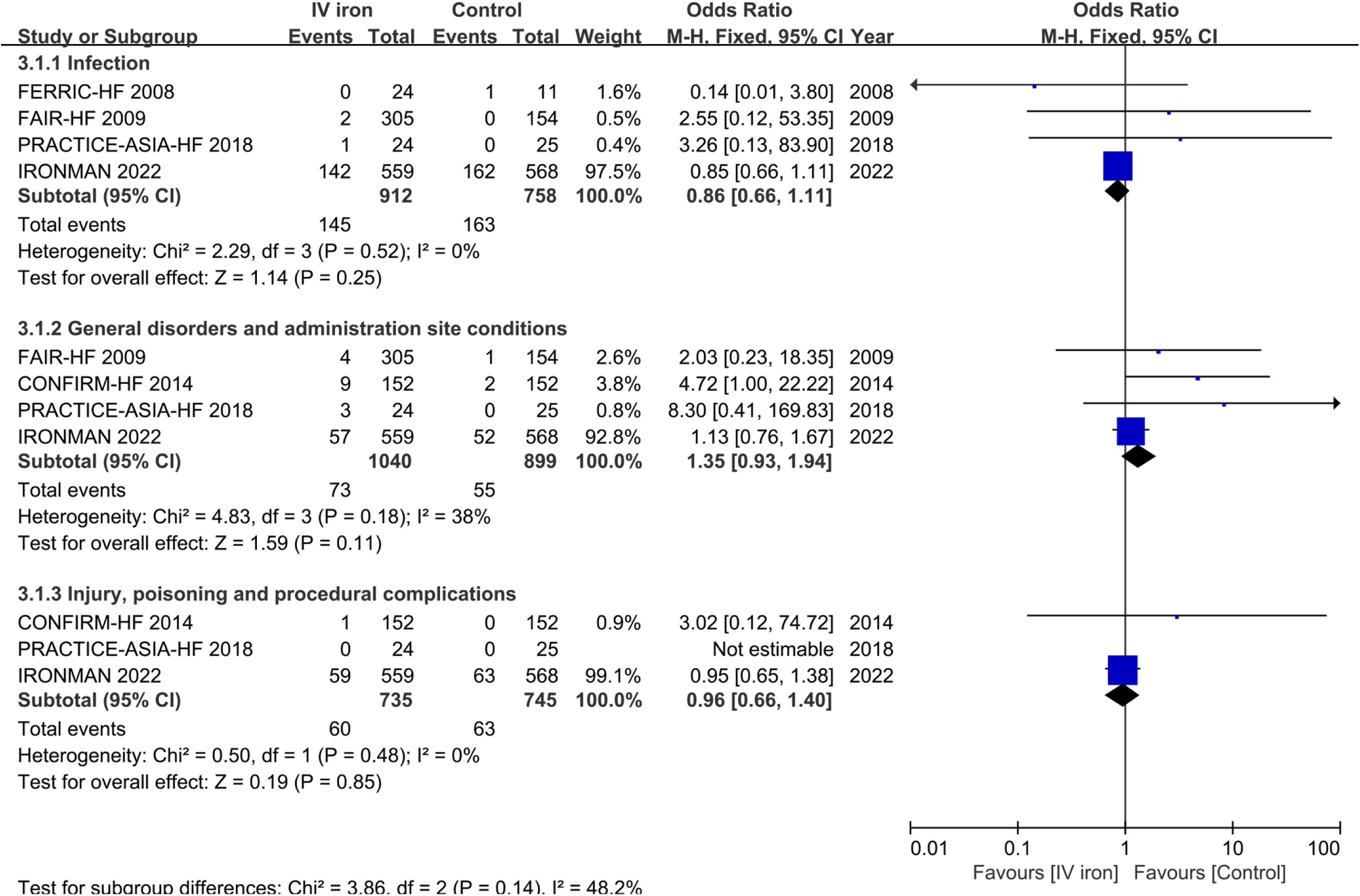
Forest plots of studies assessing adverse events in patients receiving intravenous iron therapy compared to those who received a placebo.

## DISCUSSION

This systematic review and meta-analysis revealed that intravenous iron therapy effectively reduced rehospitalization related to HF and cardiovascular causes. Furthermore, the intravenous administration of iron exhibited a potential reduction in cardiovascular mortality when evaluated alongside the hospitalization risk as a composite endpoint. The prevailing intravenous iron utilized in current trials for treating HF patients with ID was ferric carboxymaltose. Despite potential confounding factors arising from variations across studies, subgroup analyses revealed a cardiovascular protective effect, particularly in studies with longer follow-up periods and larger sample sizes. In addition, intravenous iron did not increase the incidence of adverse events. These findings thus imply that intravenous iron repletion represents a promising strategy to further mitigate residual risks associated with HF when used in conjunction with other medications for clinical management.

Despite significant advancements in therapeutic approaches for HF that greatly improve its prognosis, the recurrent hospitalization and mortality rates remain high^7, 19^. ID exerts negative impact on the prognosis of HF and poses a significant obstacle to achieving cardiovascular benefits through current treatments for HF patients ^7, 20^. The 2017 IRONOUT HF trial found that oral iron supplements had no improvement in exercise tolerance or clinical outcomes in patients with HF and ID^21–23^. In our systematic review and meta-analysis, we excluded any RCTs investigating the use of oral iron therapy because oral iron therapy is not recommended in the most recent clinical guidelines for HF and ID. In addition, including such studies would introduce unnecessary heterogeneity to our analysis and complicate the interpretation of the results^5, 20^. Both the AFFIRM-AHF trial and the IRONMAN trial demonstrated that intravenous iron derivatives effectively relieved clinical symptoms and reduced HF-related hospitalizations, with no effects on mortality^17, 22, 24^. However, the recently HEART-FID trial, the largest RCT involving patients with both HF and ID, reported that intravenous iron therapy showed no significant impact on the mortality or rehospitalizations rates^10, 15^. Our meta-analysis indicates that intravenous iron therapy was associated with substantial reductions in rates of HF hospitalizations (40%), cardiovascular death and HF hospitalizations (46%), and cardiovascular death and hospitalizations (53%). Subgroup analyses suggested that larger sample sizes and longer follow-up periods in the included studies were associated with more pronounced benefits of intravenous iron therapy. These findings aligned with a recently published meta-analysis that specifically investigated the effects of intravenous iron therapy on patients with HF^10^, but significant heterogeneities among the included studies existed. The inconsistencies observed in our analysis can be attributed to various factors, including differences in sample size, the type of HF, duration of follow-up, evaluation criteria for clinical outcomes, dosage and course of intravenous iron, co-administration of oral medications, and even the impact of the coronavirus disease 2019 pandemic^15, 17, 22, 24^. Subsequent sensitivity analyses showed a notable reduction in heterogeneities while maintaining consistent clinical outcomes. Notably, this reduction in heterogeneity was achieved by excluding the HEART-FID trial from the analysis. After conducting a comprehensive review and comparison of the HEART-FID trial with other studies, the frequent administration of sacubitril-valsartan in the HEART-FID trial potentially influenced the assessment of the positive rate of clinical events in our meta-analysis. ^15^. In the HEART-FID trial, patients with HF had a higher mean transferrin saturation at baseline compared to other studies included in our analysis, which contributed to the observed mild clinical benefit of iron supplementation in this trial ^11, 15, 22, 24, 25^. While the HEART-FID trial and AFFIRM-AHF trial used the same type of intravenous iron, it is important to highlight that there were notable differences in the dosage and timing of ferrous carboxymethylene administration between the two trials ^15, 24^. It is worth noting that a significant proportion of the heart failure population included in the HEART-FID trial was recruited during the COVID-19 pandemic. This unique circumstance may potentially impact the reporting of hospitalization rates and mortality outcomes ^15, 20^.

Iron supplementation is primarily utilized to correct cellular energetic and oxidative metabolism deficiencies related to iron deficiency, rather than serving as a therapeutic manipulation directly targeting the pathogenesis of HF ^17, 26^. Correcting iron deficiency state may enhance the efficacy of drugs that aim to reverse myocardial remodeling. One particularly noteworthy finding from our meta-analysis was the potential cardioprotective effect of intravenous iron therapy against cardiovascular death, which was highly consistent with the IRONMAN trial^22^. Our study’s conclusion was based on the assessment of composite endpoints, which may have been impacted by readmissions. Due to the unavailability of individual patient data from the eligible RCTs, our analysis was unable to directly assess the efficacy of intravenous iron repletion on cardiovascular death^20^. In addition, our data revealed a numerical benefit but did not demonstrate statistical significance in all-cause mortality. Previous studies have noted a similar trend towards improved all-cause mortality with intravenous iron supplementation, although statistical significance was not achieved. The underlying causes lie in excessive tissue iron deposition and subsequent free radical-induced tissue damage^24, 25, 27^, highlighting the need for further investigation into the optimal dosing and administration of iron therapy in HF patients. By calculating FIs and FQs in this meta-analysis, we identified a delicate result indicating that the statistically non-significant treatment effect could potentially become significant if a few specific event states were modified^18^. This suggests that the overall conclusions of our analysis may be sensitive to changes in these specific events, emphasizing the need for further investigation and cautious interpretation of the findings. These data can be interpreted in light of the definition of all-cause mortality, which encompasses a broad range of deaths unrelated to HF. The pooled estimates may be influenced by heterogeneities arising from different definitions of all-cause mortality used in the included studies. Therefore, caution is warranted when interpreting the findings in relation to all-cause mortality in this context. Therefore, whether the benefits of iron treatment for HF in improving mortality outcomes are conclusive remains uncertain.

### Limitations

The limitations still exist. Firstly, the populations included in the eligible trials exhibited heterogeneity in the type of HF, specifically in terms of acute versus chronic HF and preserved ejection fraction versus reduced ejection fraction HF ^5, 17, 28^. Due to the unavailability of individual participant data, we were unable to conduct a direct comparison of the efficacy of intravenous iron treatment in different categories of HF within our analysis.

Secondly, the discrepancies in the administration of intravenous iron among the included studies may have contributed to significant heterogeneity in our results. This variation in the dosage and administration protocols of intravenous iron treatment in HF patients underscores the need for further investigation to determine the optimal dosage and administration strategy. Thirdly, six trials included in our analysis had a recruitment size of fewer than 200 individuals with HF, which resulted in a lack of statistical power to detect significant differences^11, 29–33^; the recent large-scale trials included in our analysis enrolled a significant number of patients with COVID-19, which poses a challenge in determining whether the pandemic may have influenced our findings^15, 22, 24^. Therefore, it is crucial not to underestimate the significance of sample deviation and the potential for Type I error. In order to obtain more accurate estimates of the efficacy of intravenous iron administration, it is imperative that future RCTs prioritize comprehensive clinical designs and incorporate long-term observation of hospitalizations and mortality outcomes^34^.

## CONCLUSION

The findings from our analysis suggest that intravenous iron therapy may decrease the risk of rehospitalization for HF and result in a lower likelihood of cardiovascular death. While some controversies remain and further evidence is needed, our meta-analysis confirms that incorporating intravenous iron therapy into clinical treatment decisions for HF patients with ID could enhance current clinical practice and potentially lead to improved clinical outcomes.

## Data Availability

We extracted data from publicly available randomized controlled studies. All types of data in this manuscript are publicly available

## Acknowledgments

HW, YHL, XHG and RZ contributed to the study conception and design, and writing the manuscript. HW, YHL, JW, JYL, JJZ and JJS performed data collection and analysis. HW, YHL, XHG and RZ commented on the research design, data analysis, writing the manuscript, and supervision of the study. All authors contributed to the article and approved the submitted version.

## Supplementary Materials

Supplementary Figures 1-17

